# Blood single cell immune profiling reveals the interferon-MAPK pathway mediated adaptive immune response for COVID-19

**DOI:** 10.1101/2020.03.15.20033472

**Authors:** Lulin Huang, Yi Shi, Bo Gong, Li Jiang, Xiaoqi Liu, Jialiang Yang, Juan Tang, Chunfang You, Qi Jiang, Bo Long, Tao Zeng, Mei Luo, Fanwei Zeng, Fanxin Zeng, Shuqiang Wang, Xingxiang Yang, Zhenglin Yang

## Abstract

The coronavirus disease 2019 (COVID-19) outbreak is an ongoing global health emergence, but the pathogenesis remains unclear. We revealed blood cell immune response profiles using 5’ mRNA, TCR and BCR V(D)J transcriptome analysis with single-cell resolution. Data from 134,620 PBMCs and 83,387 TCR and 12,601 BCR clones was obtained, and 56 blood cell subtypes and 23 new cell marker genes were identified from 16 participants. The number of specific subtypes of immune cells changed significantly when compared patients with controls. Activation of the interferon-MAPK pathway is the major defense mechanism, but MAPK transcription signaling is inhibited in cured patients. TCR and BCR V(D)J recombination is highly diverse in generating different antibodies against SARS-CoV-2. Therefore, the interferon-MAPK pathway and TCR-and BCR-produced antibodies play important roles in the COVID-19 immune response. Immune deficiency or immune over-response may result in the condition of patients with COVID-19 becoming critical or severe.

---

The battles between humans and the pathogenic viruses are never ending. Epidemic diseases, including Zika (*1*), severe acute respitatory sundrome (SARS) (*2*), and Ebola (*3*), strike quickly, often killing hundreds of people during a single outbreak. On December 31, 2019, a novel coronavirus (SARS-CoV-2), belonging to the Orthocoronavirinae subfamily and distinct from middle east respiratory syndrome (MERS)-CoV and SARS-CoV, emerged in Wuhan, China (*4-6*). The rapid person-to-person spread of SARS-CoV-2, which causes the coronavirus disease 2019 (COVID-19), presents an imminent threat to public health and has caused a global health emergency (*7-9*). The World Health Organization emergency committee declared this outbreak a “public health emergency of international concern” because of its 2.1% mortality rate and person-to-person transmission (*10*). As of March 7, 2020, over 102,136 people have fallen ill with SARS-CoV-2, leading to 3,492 deaths in 93 countries, and the coronavirus continues to spread quickly all over the world. Symptoms of COVID-19 included fever, myalgia, and fatigue, as well as dry cough, shortness of breath, sputum production, headache, hemoptysis, sore throat, and diarrhea. Lymphopenia, prolonged prothrombin time, and elevated lactate dehydrogenase levels have also been observed in the patients with COVID-19 (*6*). A computed tomography (CT) can identify bilateral patchy shadows or ground glass opacity in the lungs (*6, 8, 11-13*). Despite the high infection and mortality rates, there is no specific cure for the disease because its pathogenesis remains unclear.

The human adaptive immune system plays a critical role in defending against viral infections. Many immune cells (such as leukocytes) and immune molecules (such as specific plasma proteins) are intrinsic components of blood. Therefore, the functions of blood and the immune system are inseparable. T cells and B cells are important in the pathogenesis of many—perhaps all—immune-mediated diseases (*14-17*). A T cell receptor (TCR) mediates recognition of pathogen-associated epitopes through interactions with peptide and major histocompatibility complexes (pMHCs). TCRs and B cell receptors (BCRs) are generated by genomic rearrangement at the germline level, a process termed V(D)J recombination, which has the ability to generate marked diversity among TCRs and BCRs. Parameterizing the elements of antigen-specific immune repertoires across a diverse set of epitopes may generate powerful applications in a variety of research fields for the diagnosis and treatment of infectious diseases. Here, we focus on insights into blood immune responses to COVID-19 using 5’ mRNA, TCR, and BCR V(D)J recombination analysis with single-cell resolution.

In this study, 16 participants were recruited from the Sichuan Province of China. The basic information and clinical features of the patients are listed in Table S1. The lung CT images of these subjects are shown in Fig. S1. There was one critical case (patient 1, male, 34 years old), one severe case (patient 2, male, 43 years old), and six moderate cases (patients 3-8) comprised of two males and four females aged 25-62 years old (Table S1). The two cured patients (patient 9, male, 40 years old and patient 10, female, 20 years old) were on the discharged day from hospital after they tested negative for SARS-CoV-2 and the disease signs disappeared. Three healthy people were included as normal controls (NC 1-3, Table S1). In order to differentiate the COVID-19 immune response from other infectious diseases, thereby identifying the unique COVID-19 immune response, we also recruited one case of influenza A (patient 11), one case of acute pharyngitis (patient 12), and one case of cerebral infarction (patient 13) as controls (Table S1).

The peripheral blood mononuclear cells (PBMCs) of each individual were isolated from the whole blood, and the single-cell mRNA sequencing (ScRNA-Seq) analysis of the PBMCs was performed on the 10 X genomics platform with Chromium Next GEM Single Cell V(D)J Reagent Kits v1.1 (Fig. 1A). After quality control, we obtained 134,620 PBMC cells, 83,387 TCR clones, and 12,601 BCR clones from the 16 samples. In these samples, we identified 17 completely different PBMC cell types, which could be further divided into 56 cell subtypes according to marker genes of the 134,620 cells (Fig. 1B, 1C, 1D and Table S2). The 17 cell types included CD4+ T cells identified by *LTB* and *IL7R* markers, CD8+ T cells identified by *LEF1* and *CD8A*, CD1C+_B dendritic cells identified by *BSET1* and *CD1D*, CD4+ cytotoxic T cells identified by *CST7* and *CCL4*, CD8 cytotoxic T cells identified by *CD3D* and *CD8A*, and other cell types (Fig. 1C, Table S2). This suggests that human PBMCs have evolved highly differentiated cell subtypes to resist the invasion of pathogens (*18*). There were three main states in the analysis process for reconstructing the developmental trajectories of the 56 cell subtypes, which showed a continuous trajectory of differentiating PBMCs (Fig. 1E, Fig. S2). In addition, 23 totally new candidate marker genes linked to 22 cell subtypes were identified (Fig. S3), suggesting potential roles for these newly discovered cell markers in the identification and functional study of these subtypes.

**Fig. 1.**
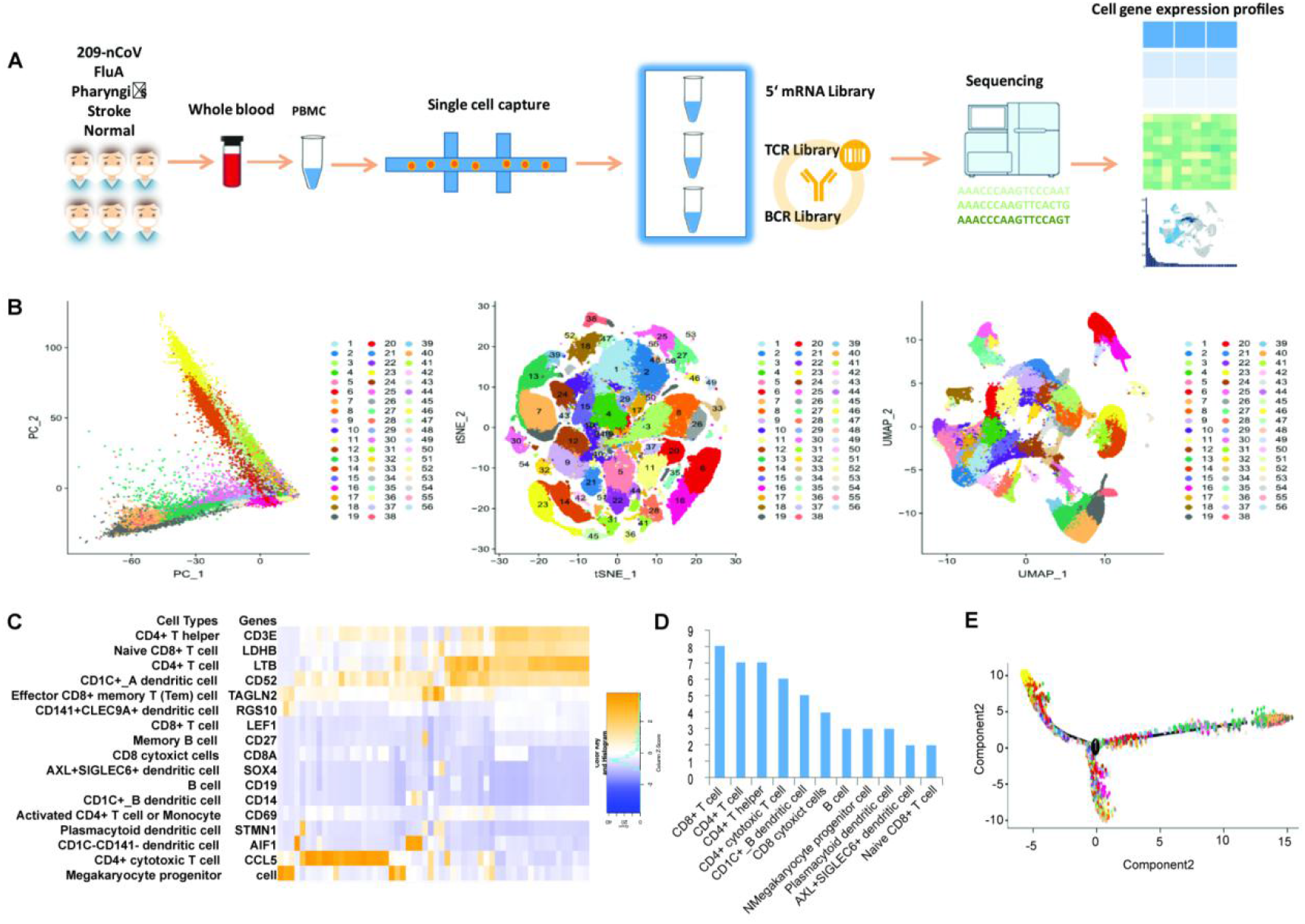
Cellular composition of the PBMCs in COVID-19 patients and controls. (A) Schematic of single-cell immune transcriptome profiles of the PBMCs in COVID-19 patients and normal controls. The PBMCs were isolated for constructing single cell 5’ mRNA, TCR, and BCR libraries using chromium single cell V(D)J v1.1 reagent kits of 10x genomics chemistry. (B) Integration analysis results of COVID-19 patients and normal controls showing principle component (PC), t-SNE algorithm, and UMAP algorithm visualization. In total, 56 cell subtypes were identified. (C) Seventeen different cell types. From these types, the 56 cell subtypes were derived. (D) Cell subtypes in each main cell type (only cell types with two or more subtypes are displayed). (E) Cell differentiation trajectory analysis, which indicates three states of cells.

To investigate the immune cell behaviors of the 56 cell subtypes in different conditions of COVID-19, we compared the cell types of the eight patients with critical, severe, or moderate COVID-19 conditions and the three normal controls (Fig. 2A). We observed that the proportion of the CD1C+_B dendritic cells (clusters 7, 13, and 30), CD8 cytotoxic T cells (cluster 9), and plasmacytoid dendritic cells (cluster 36) increased in patients with COVID-19, while the B cells (cluster 16) and CD4+ T helper cells (cluster 51)decreased in the COVID-19 patients compared to the normal controls (Fig. 2A), indicating that the number of specific clusters of immune cells adjusts to fight SARS-CoV-2 aggression.

**Fig. 2.**
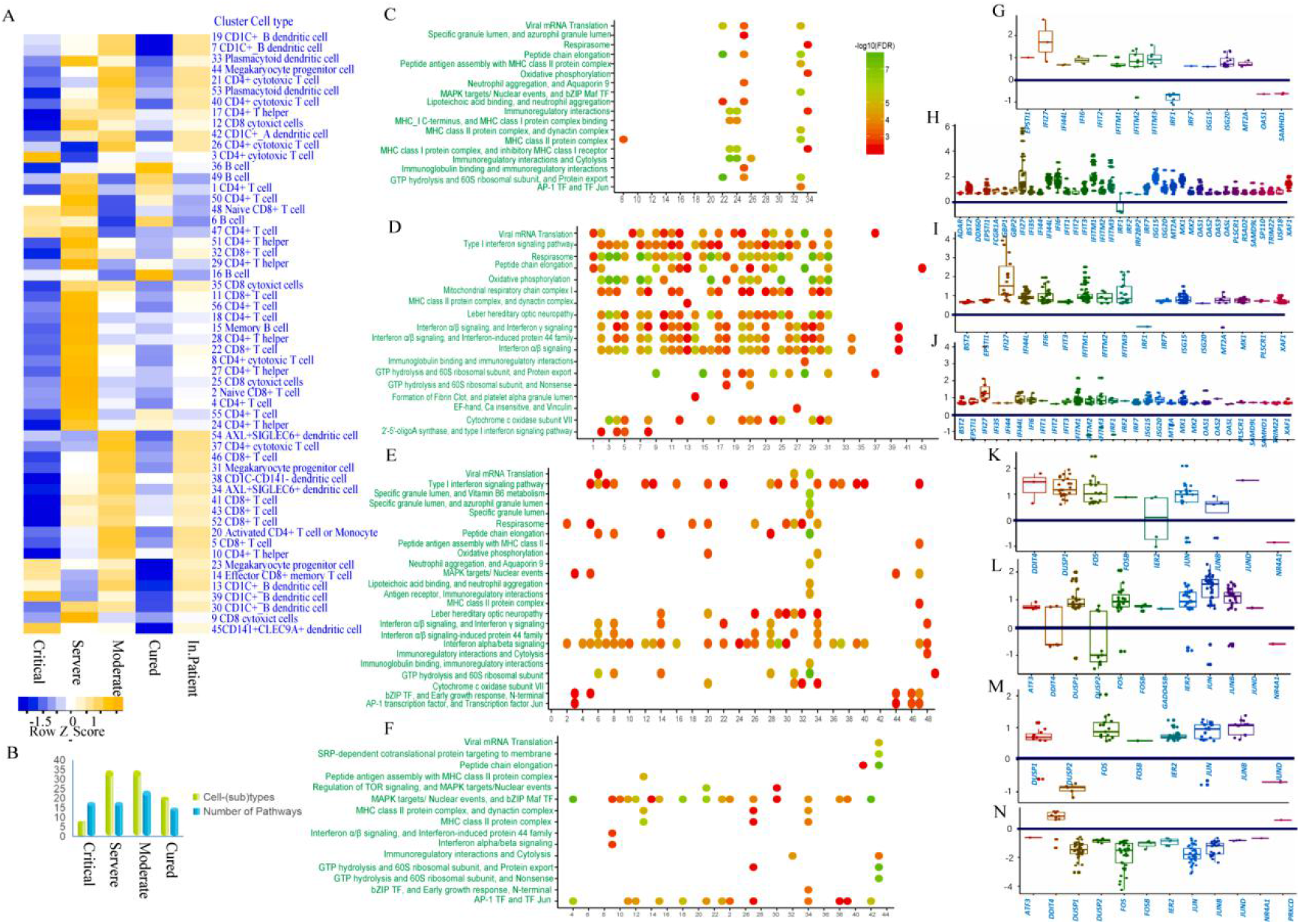
The interferon-MAPK pathway is the key response in PBMCs for SARS-CoV-2 infection. (A) Comparisons of cell type behaviors of patients with COVID-19 and normal controls. (B) Cell subtypes and the number of pathways significantly differed between patients with COVID-19 and the normal controls. (C-F) The enrichment pathways of differential expressed (DE) genes in each cell subtype. (C) The critical patient (patient 1) vs normal controls. (D) The severe patient (patient 2) vs normal controls. (E) The moderate patients (patients 3-6) vs normal controls. (F) The cured patients (patient 9 and 10) vs normal controls. (G-J) Log2 fold changes of interferon pathway genes in critical, severe, moderate, and cured patients with COVID-19 vs normal controls, respectively. Each point represents a different cell subtype. (K-N) Log2 fold changes of MAPK pathway genes in critical, severe, moderate, and cured patients with COVID-19 vs normal controls, respectively. Each point represents a different cell subtype.

To further investigate the immune cell behaviors of each cell subtype in different conditions of COVID-19, we separately compared the critical, severe, moderate, and cured patients with the normal controls (Fig. 2A). For the critical condition (patient 1), the proportion of 40 out of the 56 cell subtypes decreased; some subtypes, including CD8+ T cell (cluster 52), CD4+ T cell (cluster 55,56) and plasmacytoid dendritic cell (cluster 53), disappeared. These changes may suggest immunodeficiency in the patient. This is consistent with the lower number of lymphocytes in the patient (0.75 × 10^9^/L) compared to the normal range (1.1-3.2 × 10^9^/L, Table S1). However, some other cell subtypes, including CD8 cytotoxic T cells (cluster 9), CD1C+_B dendritic cells (cluter 7, 13, 19, 30, 39), effector CD8+ memory T (Tem) cells (cluster 14), and CD141+CLEC9A+ dendritic cells (cluster 45), increased, suggesting the limited immune response existed. In the severe condition (patient 2), the proportion of 45 of the 56 cell types increased, which may suggest over-activation of immune cells, including CD8+ T cells (cluster 1, 4, 18, 28 etc.), CD8 cytotoxic T cells (cluster 1 and cluster 25), CD4+ cytotoxic T cells (cluster 8 and cluster 21), Naive CD8+ T cell (cluster 2), plasmacytoid dendritic cell (cluster 53), and others. However, AXL+SIGLEC6+ dendritic cells (cluster 54) decreased in this patient. In the moderate condition (patients 3-8), the proportion of various cell types was between that of the critical and severe conditions. In the cured patients, effector CD8+ memory T (Tem) cells (cluster 14), megakaryocyte progenitor cells (cluster 23), CD1C+_B dendritic cells (cluster 39), and CD141+CLEC9A+ dendritic cells (cluster 45) decreased significantly. These results suggested a wide range of immune responses involved in the blood circulation system after SARS-CoV-2 infection.

Then, we analyzed the gene expression differences in each cell subtype of the COVID-19 conditions and controls and identified a number of differentially expressed (DE) genes (External Databases S1-S4). To further explore the enriched signal pathways of the DE genes in each cell subtype, we analyzed the enriched pathways of the DE genes in each cell cluster using local string network analysis. In patients with critical, severe, moderate, or cured conditions, we found that 18 (in eight cell subtypes), 18 (in 35 cell subtypes), 24 (in 35 cell subtypes), and 15 (in 21 cell subtypes) signal pathways, respectively, were over-activated or inactivated for each disease condition (Fig. 2B-F, Table S3-6). These significantly enriched signal pathways were mainly involved in viral mRNA translation (high gene expressions in AXL+SIGLEC6+ dendritic cells, CD4+ T cells, and other cells) (Fig. S4-5), interaction alpha/beta signaling (Fig. S6-7), a mitogen-activated protein kinase (MAPK) pathway (Fig. S8-9), immunology interactions between lymphoid and non-lymphoid cells (CD1C+_B dendritic cells and CD4+ cytotoxic T cells) (Fig. S10-11), and major histocompatibility complex (MHC) class II protein complex (B cells and CD1C+_B dendritic cells) (Fig. S12-13), suggesting that antigen presentation for targeting the virus was activated.

In the severe condition (patient 2), 18 signaling pathways were significantly changed in more than half of the cell subtypes compared to the normal controls. Among these pathways, the most important was over-activation of the interferon-related signaling pathways, including 2’-5’-oligoadenylate synthase and type I interferon signaling pathway(naive CD8+ T cells, CD4+ T cells, and other cells) and interferon alpha/beta/gamma signaling (naive CD8+ T cells, CD4+ T cells, and other cells) (Fig. 2C, Table S3). In other COVID-19 conditions, the interferon signal was also over-activated to varying degrees. However, in the cured patients, the interferon signal was just mildly activated compared to the patients with severe and moderate conditions. In the cured condition, the signal pathway related to mRNA translation of the virus was suppressed, while the signal pathway related to MAPK was significantly enriched (Fig. 2C, Table S3-6). To detect whether these signaling pathway changes were specific to COVID-19, we compared the COVID-19 patients with the patients with influenza A (patient 11), acute pharyngitis (patient 12), and cerebral infarction (patient 13) and found that the signaling pathways were still enriched after filtering with these related disease controls (Fig. S14). Among these pathways, the interferon-MAPK signaling pathway was the major blood immune response for COVID-19 infection.

The activation of the MAPK signaling pathway by interferon involves a set of defense mechanisms evolved by human beings to fight against viral infections (*19-21*). To further explore the expression differences of genes involved in different cell types in the various COVID-19 conditions, we analyzed the gene expressions in patients with COVID-19 and compared them to those of the controls (Fig. 2G-J, Fig. S7). First, we observed up-regulation expression of the interferon signal genes in the COVID-19 patients. Among them, interferon α-inducible protein 27 (IFI27), originally known to involve in innate immunity and to intervene in cell proliferation (*22*), was increasingly expressed in many cell types, including CD1C+_B dendritic cells, CD1C-CD141-dendritic cells, CD141+CLEC9A+ dendritic cells, CD8 cytotoxic T cells, and plasmacytoid dendritic cells. Second, the expressions of *IFITM1, IFITM3, and IFI6*, which were shown to be related to interferon-related pathway and associated with [Influenza](http://www.malacards.org/card/influenza) and [West Nile Virus](https://www.malacards.org/card/west_nile_virus) (*23-26*), were strikingly up-regulated in all of these cells. The expression trends of these genes were similar in the four COVID-19 conditions. The results indicate that interferon signals, especially interferon alfa/beta/gamma and interferon type I, are activated in the blood of patients with COVID-19.

Then, in the downstream of the interferon signaling, we found that MAPK signaling pathway transcription factors were activated in hospitalized patients with COVID-19, including high expressions of *FOS, JUN, JUNB*, and *DUSP1* (Fig. 2K-N, Table S8). These genes were previously shown to regulate [cell invasion, migration](https://www.ncbi.nlm.nih.gov/pubmed/26754630/), [proliferation](https://pubmed.ncbi.nlm.nih.gov/27890927/) and innate immunity by inhibiting pro-inflammatory cytokine production (*27, 28*). In three groups of inpatients with different conditions, the up-regulated expression of these genes was observed in CD1C+_B dendritic cell, CD1C-CD141-dendritic cell, CD141+CLEC9A+ dendritic cell, CD8 cytotoxic T cells, and plasmacytoid dendritic cells. Interestingly, in contrast to the three groups of inpatients, the expression pattern of these genes in the two cured patients was the opposite. In the cured patients, down-regulated expression was detected, indicating that the MAPK signaling pathway was inhibited in these cells. These results indicate that down-regulation of the MAPK signal pathway may be one sign of patient recovery.

To further explore the antibody clonal proliferation of TCR and BCR in COVID-19, we conducted TCR and BCR V(D)J single cell transcriptome analysis. Using integration analysis, we detected 83,387 TCR cell clones and 12,601 BCR clones in the 16 subjects (Fig. 3A). This result revealed the antibody repertoire in patients with COVID-19 and showed huge diversities of TCR and BCR clones in each individual. In the critical condition (patient 1), the fewest TCR clones (1,232) were detected, while, in the severe condition (patient 2), more TCR clones (7,161) were detected. In one moderate patient (patient 8), the most TCR clones (20,370) were detected. According to these findings, the fewer the TCR clones, the more severe the COVID-19 condition. The distribution of all TCR clones and the number of TCR clones of each patient are shown in Fig. 3B and Table S9. Using evolutionary analysis of the top TCR antibody sequences revealed by TCR sequencing, we found that TCRα sequences of CASSEGVGTPFDEQFF (patient 2), CASSLGLAGDLDEQFF and CASNQGLAGGRLYNEQFF (patient 4), CASSQERGVYNEQFF (patient 6) and CASSEVWASDHEQYF (patient 10) were closely related (Fig. S15), suggesting that these antibody sequences may have a special activity for SARS-CoV-2 (and influenza A). Compared to TCR, the number of BCR clones detected was much lower (Fig. 3C, Table S10). However, very strong BCR antibodies were not induced in most of the patients except in two moderate patients (patients 6 and 8).

**Fig. 3.**
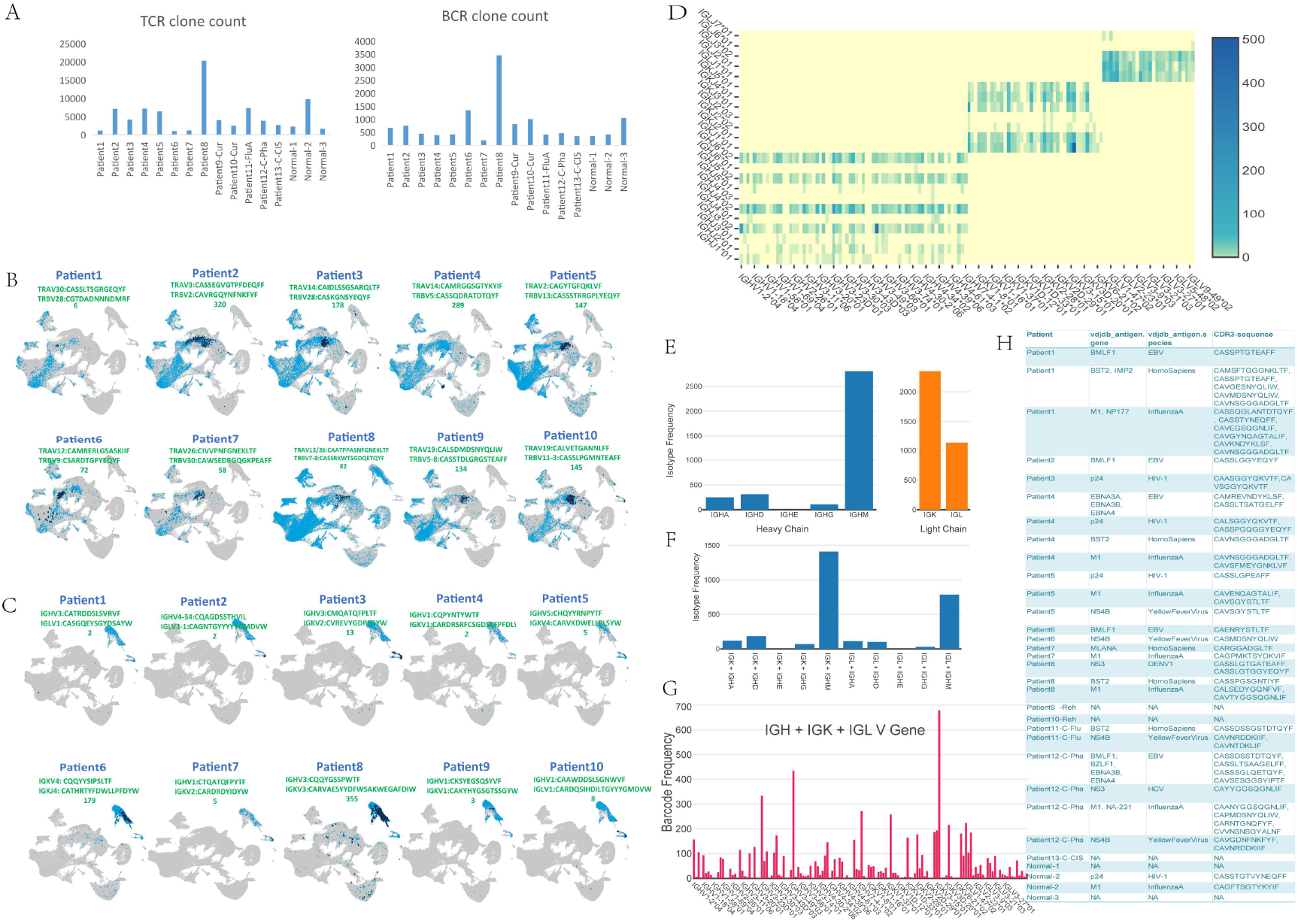
TCR and BCR V(D)J clone expression in patients with COVID-19. (A) The figure on the left shows the number of TCR clones detected by V(D)J in each patient, and the figure on the right shows the number of BCR clones detected by V(D)J in each patient. (B) The distribution of TCR clones in cell clusters of patients with COVID-19. The light blue dots indicate the distribution of all TRAV and TRBV clones, and the dark blue dots indicate the antibody sequence and quantity of the clone with the strongest TCR clone signal of the patient 8 (moderate condition). (C) The distribution of BCR clones in the cell clusters of each patient. Among them, light blue dots indicate the distribution of total IGHV, IGLV, and IGKV in this patient. The dark blue dots indicate the clones with the strongest signals in this patient. Patient 9 has the strongest B cell antibodies among all COVID-19 patients. (D) V-J heatmap of IGH + IGK + IGL in the B cells of patient 9. (E) The left and right graphs represent the isotype frequency of the heavy chain and the light chain detected in the B cells of patient 9, respectively. (F) The distribution of paired isotypes in the B cells of patient 9. Abscissa includes different chains, and the ordinate is frequency. (G) The usage of the V gene in the B cells of patient 9. (H) List of the known antigens and antibodies in patients with COVID-19.

In one moderate patient (patient 9) (Fig. 3D-G), we detected 361 clones of CDR3 IGLorK/IGH sequences of CQQYGSSPWTF/CARVAESYYDFWSAKWEGAFDIW and 119 clones of CQQYNSYSF/CARSGPITIFGVVIKGRGGDAFDIW (Fig. 3C, Table S10). In another moderate patient (patient 7), four high frequency clones were identified, including CQQYYSIPSLTF/CATHRTYFDWLLPFDYW (179) (Fig. 3C, Table S10). These results suggest that the antibodies produced by patients 7 and 9 may have a SARS-CoV-2 specific affinity.

We then analyzed top TCR encoded antibody sequences for known antigens, and some of them were annotated in the VDJdb antigen categories database (Fig. 3H). In patients with COVID-19, we detected immune responses to known antigens, including Epstein-Barr virus (EBV), HIV-1, influenza A, yellow fever, and other RNA viruses, as well as DNA viruses such as cytomegalovirus (CMV). In addition, autoantigens were detected in the severe condition (patient 2) and the moderate condition (patients 4, 8, and 9). In patient 2, the autoantigen response was obvious activation, which indicated that this severe disease condition may be related to strong autoimmunity. On the contrary, in the critical condition (patient 1), only two antibodies against the Ebola EBV antigen were detected. These results further suggest that the critical condition may be caused by insufficient immunity, while the severe condition may be aggravated by excessive autoimmunity. In the normal condition, no immune response to these antigens was detected for NC 1 and NC 3; for the NC 2, two antigens were detected (HIV-1 and influenza A) (Fig. 3H). For the patient with influenza A (patient 11), the antigen and autoimmune responses of the yellow fever virus were detected. For the patient with acute pharyngitis (patient 12), EBV, HIV-1, influenza A and yellow fever virus antigens were detected, but there was no autoimmune response. For the patient with cerebral infarction (patient 13), no known antigens were detected. These results indicate that humans might use known immune recognition (mainly known RNA viruses) to fight against SARS-CoV-2.

Type I interferon (IFN-I) is crucial for promoting antiviral defenses through the induction of antiviral effectors (*29*). In this study, our single cell transcriptome results revealed that a set of genes involved in the interferon-MAPK pathway were up-regulated in the COVID-19 patients. To further validate this finding, we performed quantitative real-time reverse transcriptase polymerase chain reaction (RT-PCR) testing to detect the expressions of *IRF27, BST2*, and *FOS* in another cohort of three critical, three server, 19 moderate, three mild and 10 cured patients with COVID-19 and five normal controls (Fig. 4A). *IIF27*, an immune biomarker which was reported to be related to [cell proliferation and invasion](https://pubmed.ncbi.nlm.nih.gov/29580248/) and could discriminate between influenza and bacteria in patients with suspected respiratory infection (*30*), showed the most strongest up-regulation in COVID-2019 patients’ PBMCs in the interferon signaling pathway in the single cell transcriptome data. The real-time PCR results showed that *IRF27* was up-regulated 8.1 times more in critical patients, 51.7 times in severe patients, 39.9 times in moderate patients, 38.6 times in mild patients, and 29.5 times in cured patients compared to the normal controls (Fig. 4A). These results suggest that *IRF27* is a candidate marker gene for SARS-CoV-2 infection. *BST2*, which was previously shown to play a role in pre-B-cell growth (*31*), also showed up-expression in COVID-19 patients. Consistent with our single cell transcription data, the expression profile of *FOS*, which is a transcription factor mediating MAPK pathway signaling, showed up-regulation in COVID-19 patients but down-regulation in cured patients. This result suggests that FOS is a candidate marker gene for cured COVID-19 patients. We further performed PBMC immunofluorescence testing to compare the protein expressions of IFI27 between patients with COVID-19 and controls. Immunofluorescence staining showed that IFI27 was highly expressed in the whole lymphocyte of patients with COVID-19, but only partially expressed on the lymphocyte of the controls (Fig. 4B).

**Fig. 4.**
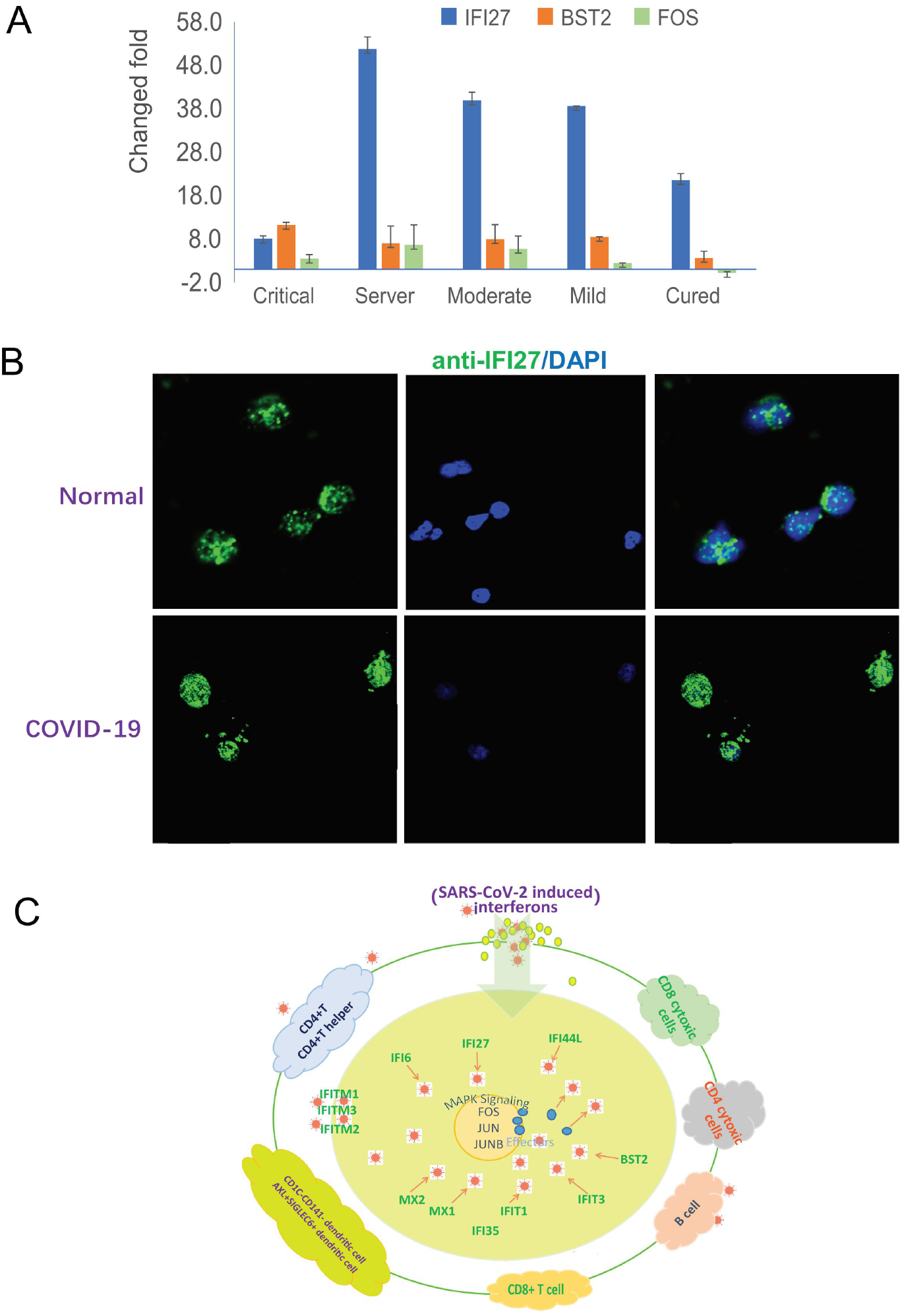
The interferon-MAPK pathway in response to SARS-CoV-2 infection. (A) Real-time PCR validation of *IFI27* and *BST2* in the interferon pathway and FOS in the MAPK pathway. *IFI27* and *BST2* are up-regulated in patients with COVID-19. FOS is up-regulated in hospitalized patients but down-regulated in cured patients. (B) Immunofluorescence staining of IFI27 in PBMCs of patients with COVID-19 and normal controls. (C) Anti-SARS-CoV-2 response of the blood system. After the virus reaches the blood immune cells, it can activate multiple cell subtypes of the interferon signal pathway to produce effectors, such as IFI27, IFITM1, and IFITM3, to fight the virus. Through downstream activation of MAPK, the expression of downstream effectors is activated by key transcription factors, FOS, JUN, and JUNB, which lead to a wide range of antiviral responses in the blood system.

In summary, we demonstrated that SARS-CoV-2 can enter the blood through the circulatory system and cause a blood immune response after infection through the respiratory system. Therefore, in patients with COVID-19, the damage is related to multiple organs (*32*). The leukocyte immune responses involve many cell subtypes, including CD8+ T cells, CD8 cytotoxic T cells, CD4+ T cells, CD4+ cytotoxic T cells, CD1C+_B dendritic cells and other cell subtypes. After the virus reaches the blood immune cells, it might replicate-mainly in AXL+SIGLEC6+ dendritic cells, CD4+ T cells, and naive CD8+ T cells and might activate multiple cell subtypes of the interferon signal pathway to produce effectors, such as IFI27, IFITM1 and IFITM3, to fight the SARS-CoV-2 virus. The similar process also happened to [Influenza](http://www.malacards.org/card/influenza) and [West](https://www.malacards.org/card/west_nile_virus) Nile Virus infection in human (*23, 24*). Through downstream activation of MAPK, the expression of downstream effectors is activated by key transcription factors, such as *FOS, JUN*, and *JUNB*, which lead to a wide range of antiviral responses in the blood system (Fig. 4C). These results suggested that the immune response may come back the normal situation when the patients recovery, and the MAPK signal is thus inhibited.

In conclusion, in this study, we revealed the immune response process of SARS-CoV-2 entering the blood circulation system using immune profiling analysis with single-cell resolution. In the critical condition, the number of different cell types, expressed genes, and TCR/BCR responses indicated a relationship between the critical nature of the patient’s condition and his immunodeficiency. On the contrary, the severe condition was shown to be related to a strong autoimmune response and over-activated interferon pathway. These results suggested that immune deficiency or immune over-reacts may make patients with COVID-19 go to worse condition because of their imbalance adaptive immune response (*33-35*). Our study also shows that the activation of the interferon-MAPK signaling pathway and TCR- and BCR-produced antibodies play key roles in combating SARS-CoV-2 infection. These results will provide important information for COVID-19 treatment and drug development.

## Data Availability

All sequencing data will be deposited in a public database and can be accessed online through a web portal.

## Acknowledgments

We thank all participants for supporting in this study. Furthermore, we gratefully acknowledge the patients and control subjects for their donation of peripheral blood samples. We thank Genergy Bio for support with V(D)J sample processing and sequencing library preparation. We thank Y.B. Mei for helpful discussions.

## Funding

This work was supported by the Sichuan Science and Technology Program (2020YFS0014 to Z.Y.), the Chinese Academy of Medical Sciences (No.2019-I2M-5-032), the National Key Research and Development Program of China (2016YFC20160905200), and the National Natural Science Foundation of China (81790643).

## Author contributions

Z.Y. Y.S., B.G. and L.J. applied for research funds; Z.Y. and L.H. designed the experiments; L.H., Y.S., B.G. L.J., J.T., C.Y., Q.J, B.L., T.Z, M.L., S.W., and X.Y. recruited participants. L.H. performed the PBMC isolation experiments and analyzed the data; X.L. performed the real-time PCR analysis; J.Y. performed the immunofluorescence experiment; L.H. and Z.Y. wrote the manuscript. All authors read and approved the manuscript.

## Competing interests

The authors have no competing interests to declare.

## Supplementary Materials

Fig. S1 CT images of the lungs of the 10 COVID-19 patients. V(D)J analysis showed varying degrees of ground glass-like changes in the lungs.

Fig. S2 Trajectory of differentiating PBMCs of 56 cell subtypes.

Fig. S3 Violin map and distributed cell (sub)types of 23 newly identified marker genes.

Fig. S4 Differential expression patterns of viral mRNA translation genes in COVID-19 patients vs controls.

A. Critical vs normal

B. Severe vs normal

C. Moderate vs normal

D. Cured vs normal

E. SARS-COV-2 vs normal

F. SARS-COV-2 vs influenza A

G. SARS-COV-2 vs acute pharyngitis

H. SARS-COV-2 vs cerebral infarction

Fig. S5 viral mRNA gene expression in 56 cell types

Fig. S6 Differential expression patterns of interaction alpha/beta signaling genes in COVID-19 patients vs other diseases

A. SARS-COV-2 vs influenza A

B. SARS-COV-2 vs acute pharyngitis

C. SARS-COV-2 vs cerebral infarction

Fig. S7 mRNA gene expression in 56 cell types of interaction alpha/beta signaling

Fig. S8 Differential expression patterns of MAPK signaling genes in COVID-19 patients vs other diseases

A. SARS-COV-2 vs influenza A

B. SARS-COV-2 vs acute pharyngitis

C. SARS-COV-2 vs cerebral infarction

Fig. S9 mRNA gene expression in 56 cell types of MAPK pathways

Fig. S10 Differential expression patterns of immunoregulatory interactions between a lymphoid cell and a non-lymphoid cell and cytolysis genes in COVID-19 patients vs controls

A. Critical vs normal

B. Severe vs normal

C. Moderate vs normal

D. Cured vs normal

E. SARS-COV-2 vs normal

F. SARS-COV-2 vs influenza A

G. SARS-COV-2 vs acute pharyngitis

H. SARS-COV-2 vs cerebral infarction

Fig. S11 mRNA gene expression in 56 cell types of immunoregulatory interactions between a lymphoid cell and a non-lymphoid cell and cytolysis genes

Fig. S12 Differential expression patterns of MHC class genes in COVID-19 patients vs controls

A. Critical vs normal

B. Severe vs normal

C. Moderate vs normal

D. Cured vs normal

E. SARS-COV-2 vs normal

F. SARS-COV-2 vs influenza A

G. SARS-COV-2 vs acute pharyngitis

H. SARS-COV-2 vs cerebral infarction

Fig. S13 MHC class gene expression in 56 cell types

Fig. S14 Enriched pathways in COVID-19 vs other disease conditions

A. SARS-COV-2 vs influenza A

B. SARS-COV-2 vs acute pharyngitis

C. SARS-COV-2 vs cerebral infarction

Fig. S15 Evolutionary conservation analysis of top TCR- and BCR-produced antibodies

A. TCRA

B. TCRB

C. IGH

D. LGL

E. IGK

Table S1 Clinical and blood routine test information of all participants, analyzed using V(D)J transcriptome analysis

Table S2 List of known marker genes, new marker genes, and highly expressed genes of 56 cell (sub) groups, identified by integrated analysis of all sequenced samples

Table S3 Significant enriched pathways of each cell (sub)type in a critical patient compared with the normal controls

Table S4 Significant enriched pathways of each cell (sub)type in a severe patient compared with the normal controls

Table S5 Significant enriched pathways of each cell (sub)type in moderate patients compared with the normal controls

Table S6 Significant enriched pathways of each cell (sub)type in cured patients compared with the normal controls

Table S7 The differential expression of interferon-related genes in each cell (sub)type of the COVID-19 patients compared with the normal controls

Table S8 The differential expression of MAPK signal genes in each cell (sub)type of the COVID-19 patients compared with the normal controls

Table S9 List of the most enriched clone antibodies in each patient using TCR single cell transcriptome analysis

Table S10 List of the most enriched clone antibodies in each patient using BCR single cell transcriptome analysis

External Database S1 Differential expressed genes in each cell subtype in a COVID-19 critical patient and the normal controls

External Database S2 Differential expressed genes in each cell subtype between a COVID-19 severe patient and the normal controls

External Database S3 Differential expressed genes in each cell subtype between COVID-19 moderate patients and the normal controls

External Database S4 Differential expressed genes in each cell subtype between COVID-19 cured patients and the normal controls

## Notes

### Competing Interest Statement

The authors have declared no competing interest.

